# Post COVID-19 condition of the Omicron variant of SARS-CoV-2

**DOI:** 10.1101/2022.05.12.22274990

**Authors:** Shinichiro Morioka, Shinya Tsuzuki, Michiyo Suzuki, Mari Terada, Masako Akashi, Yasuyo Osanai, Chika Kuge, Mio Sanada, Keiko Tanaka, Taketomo Maruki, Kozue Takahashi, Sho Saito, Kayoko Hayakawa, Katsuji Teruya, Masayuki Hojo, Norio Ohmagari

**Author notes:** Corresponding author: Shinichiro Morioka, Address: 1-21-1 Toyama, Shinjuku-ku, Tokyo 162-8655, Japan.

## Abstract

**Background:** No epidemiological data on post coronavirus disease (COVID-19) condition due to Omicron variant has been reported yet.

**Methods:** This was as a single-center, cross-sectional study, that interviewed via telephone the patients who recovered from Omicron COVID-19 infection (Omicron group), and surveyed via self-reporting questionnaire those patients infected with other strains (control group). Data on patients’ characteristics, information regarding the acute-phase COVID-19, as well as presence and duration of COVID-19-related symptoms were obtained. Post COVID-19 condition in this study was defined as a symptom that lasted at least 2 months within 3 months since the onset of COVID-19. We investigated and compared the prevalence of post COVID-19 condition in both groups after performing propensity score matching.

**Results:** We conducted interviews for 53 out of 128 patients with Omicron, and obtained 502 responses in the control group. After matching, 18 patients each in Omicron and control group had improved covariate balance of the older adult, female sex, obese patients, and vaccination status. There were no significant differences in the prevalence of each post-acute COVID-19 symptoms between the two groups. The numbers of patients with at least one post-acute COVID-19 symptom in the Omicron and the control group were 1 (5.6%) and 10 (55.6%) (p=0.003), respectively.

**Conclusion:** The prevalence of post Omicron COVID-19 conditions was less than that of the other strains. Further research with more participants is needed to investigate the precise epidemiology of post COVID-19 condition of Omicron, and its impact on health-related quality of life and social productivity.

## Introduction

Coronavirus disease (COVID-19) has become a global pandemic with 513 million cumulative cases and 6.2 million deaths worldwide as of May 4, 2022 [1]. Following the onset of COVID-19, 49% of patients had at least one prolonged symptom, lasting more than 12 months [2]. These post-acute sequelae of severe acute respiratory syndrome coronavirus 2 (SARS-CoV-2) infection are known as the post COVID-19 condition or long COVID[3][4]. Studies have shown that the common symptoms of the post COVID-19 condition included fatigue, shortness of breath, dysosmia, hair loss and cognitive impairment, and multiple symptoms often overlap and persist [2, 5, 6].

A prospective longitudinal observational study revealed the difference in the prevalence of acute-phase symptoms in patients with Omicron and Delta variants of SARS-CoV-2 [7]. In particular, sore throat was more common in Omicron than in Delta, while loss of smell was infrequent in Omicron than Delta. However, no epidemiological data on post COVID-19 condition of Omicron has been reported yet. Considering that many patients were infected with Omicron variant worldwide [8][9], several patients may suffer from post COVID-19 condition due to Omicron. In this study, we aimed to investigate and compare the prevalence of the post COVID-19 condition of omicron with that of the other strains.

## Methods

This study was a single-center, cross-sectional study, using telephone interviews and a self-reporting questionnaire survey for the omicron and control group, respectively. The patients in telephone and self-reported survey provided an oral and written informed consent, respectively. This study was reviewed and approved by the ethics committee of the Center Hospital of the National Center for Global Health and Medicine (NCGM) (NCGM-G-004406-00, NCGM-G-004439-00).

## Study participants and study period

### Omicron group

Patients who were admitted due to COVID-19 at the National Center for Global Health and Medicine (NCGM) between December 1, 2021 and February 9, 2022 were invited for telephone interviews. The patients were positive for SARS-CoV-2 E484A mutation, and negative for L452R mutation, as confirmed by the reverse transcription-polymerase chain reaction (rtPCR).

Patients who died during admission were excluded from the study. The telephone interviews were conducted from April 14, 2022 to April 26, 2022 after confirming that 3 months had passed since the onset of COVID-19 symptoms for each patient. The investigators (M Suzuki, M Sanada, MA, YO, MT, KT, CK, TM) administered a one-on-one structured telephone interviews with the patients using an interview guide (Appendix 1). The interview lasted for 5-10 minutes, and each patient was interviewed only once. The investigators were nurses, researchers, and physicians working in the NCGM. In the individual telephone interviews, the participants were asked regarding their symptoms and the duration, and were recorded by checking the electronic files. If the participants were unable to recall the symptom duration, the answers were regarded as missing values.

### Control group

The participants recruited in the control group were patients who recovered from COVID-19, and who visited the outpatient service of the Disease Control and Prevention Center in the NCGM between February 2020 and November 2021 to undergo a pre-donation screening test for COVID-19 convalescent plasmapheresis [6]. All participants in this study were Japanese since the screening test was designed only for Japanese patients. Most of the participants had received acute-phase treatment for COVID-19 in other hospitals. A self-reporting, online or paper-based questionnaire was sent to eligible patients in February 2022, without any reminders (Appendix 2). Participation in this survey was voluntary, but not anonymous. The participants were requested to complete and return the questionnaire.

We developed the questionnaire based on previous similar studies [2, 5, 6, 10-15], including findings from our previous work on prolonged and late-onset symptoms of COVID-19 [16]. We attempted to minimize the number of questions required to maximize the response rate. Six non-medical employees in NCGM were included in the pilot testing. They provided feedback on the content, clarity, and format of the items, as well as whether the survey questions were self-explanatory. Revisions were made on the questionnaire according to their feedback.

### Items investigated

Information obtained included age, sex, ethnicity, smoking history, body mass index (BMI), underlying medical conditions, COVID-19 vaccination status, details on acute phase of COVID-19, as well as presence and duration of symptoms of post COVID-19 condition. Disease severity based on previous literature [2, 12] was categorized as follows: 1) mild, no oxygen therapy; 2) moderate, oxygen therapy without mechanical ventilation; and 3) severe, mechanical ventilation with or without extracorporeal membrane oxygenation.

### Definition of post COVID-19 condition

The definition of post COVID-19 condition in this study was a symptom that persisted for at least 2 months within 3 months since the onset of COVID-19. The post COVID-19 condition included fatigue, shortness of breath, cough, dysosmia (including anosmia), dysgeusia (including ageusia), hair loss, depressed mood, brain fog, loss of concentration, and memory disturbance. Since this study did not include patient consultations, it could not be ascertained whether the symptoms were due to COVID-19 or an alternative diagnosis [3].

### Statistical analysis

The patient characteristics, presence of pneumonia, disease severity, and treatment in the acute phase of COVID-19 were expressed as median and interquartile range (IQR) for continuous variables, and as absolute values (n) with % for categorical variables.

A multivariable logistic regression model was developed to estimate a propensity score for being the Omicron group. Age (older adult), sex (female), BMI ≥ 25 and vaccination status (vaccinated at least twice at the time of the survey) were included in the model as the four factors were associated with increased risk of post COVID-19 condition [17, 18]. Propensity score matching (PSM) was performed using the nearest neighbor matching with a caliper width of 0.2 [19]. The standardized difference was used to measure covariate balance, and an absolute standardized difference above 10% was interpreted as a meaningful imbalance. Subsequently, the proportion of patients with prolonged symptoms, and those with symptoms lasting at least 2 months within 3 months since the onset have been described among the matched population.

The level of significance in all statistical tests was set at α = 0.05. Data were analyzed using SPSS^®^ Statistics version 25.0 software (IBM^®^, Armonk, NY, USA) and R, version 4.1.3 (R Foundation for Statistical Computing; 2018, Vienna, Austria).

## Results

### Omicron group

A total of 128 patients were potentially eligible for telephone interviews. Of whom, one died after discharge. Four opted not to participate, while interviews could not be done in 22 patients due to medical condition such as dementia, and in one patient due to language barrier. Meanwhile, 47 patients could not be contacted via telephone. Overall, 53 patients were included and completed the interviews.

### Control group

A self-reporting questionnaire was sent to 1148 patients (958 online; patients; 190 paper-based) who had recovered from COVID-19. Of which, 502 responses (413 online;89 paper-based) were obtained. The overall response rate was 43.7% (43.1% online; 46.8% paper-based). Among the 502 patients, 133 (31.5%), 205 (48.6%), and 84 (19.9%) (80 missing) got infected with COVID-19 between February 2020 and October 2020, November 2020 and June 2021 (Alpha strains predominant), and July 2021 and October 2021 (Delta strains predominant), respectively[8][20].

### Characteristics of the participants in the Omicron and the control group before and after PSM

The characteristics of the participants in the Omicron and the control group before and after PSM are summarized in **Table 1** and **Table 2**, respectively. Before matching, the mean ages (years, interquartile range: IQR) and mean BMI (IQR) in the Omicron and the control group were 56.0 (35.0, 69.5) and 48.0 (42.0, 55.0), and 24.4 (22.2, 26.8) and 23.1 (20.7, 25.9), respectively. Fourteen patients (26.4%) in the Omicron group and 300 patients (59.8%) in the control group were women. Forty-five patients (84.9%) in the Omicron group and 11 patients (2.2%) in the control group tested positive for SARS-CoV-2 at least seven days after their second COVID-19 vaccination when immunity had developed [21]. Forty-two patients (79.2%) in the Omicron group and 393 patients (86.4%) in the control group had mild disease. Missing values ranged from 0% to 15.5%.

**Table 1.**
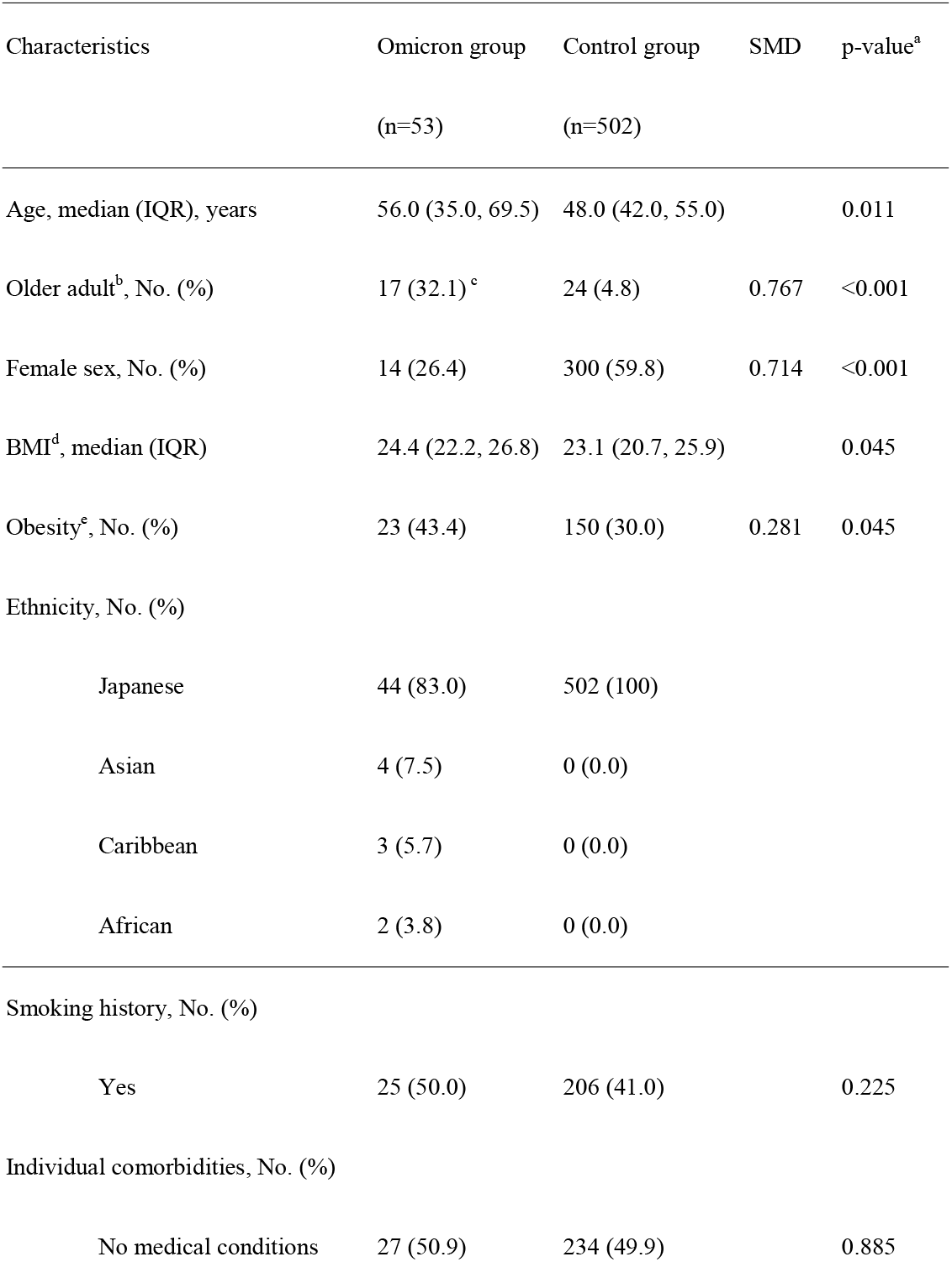
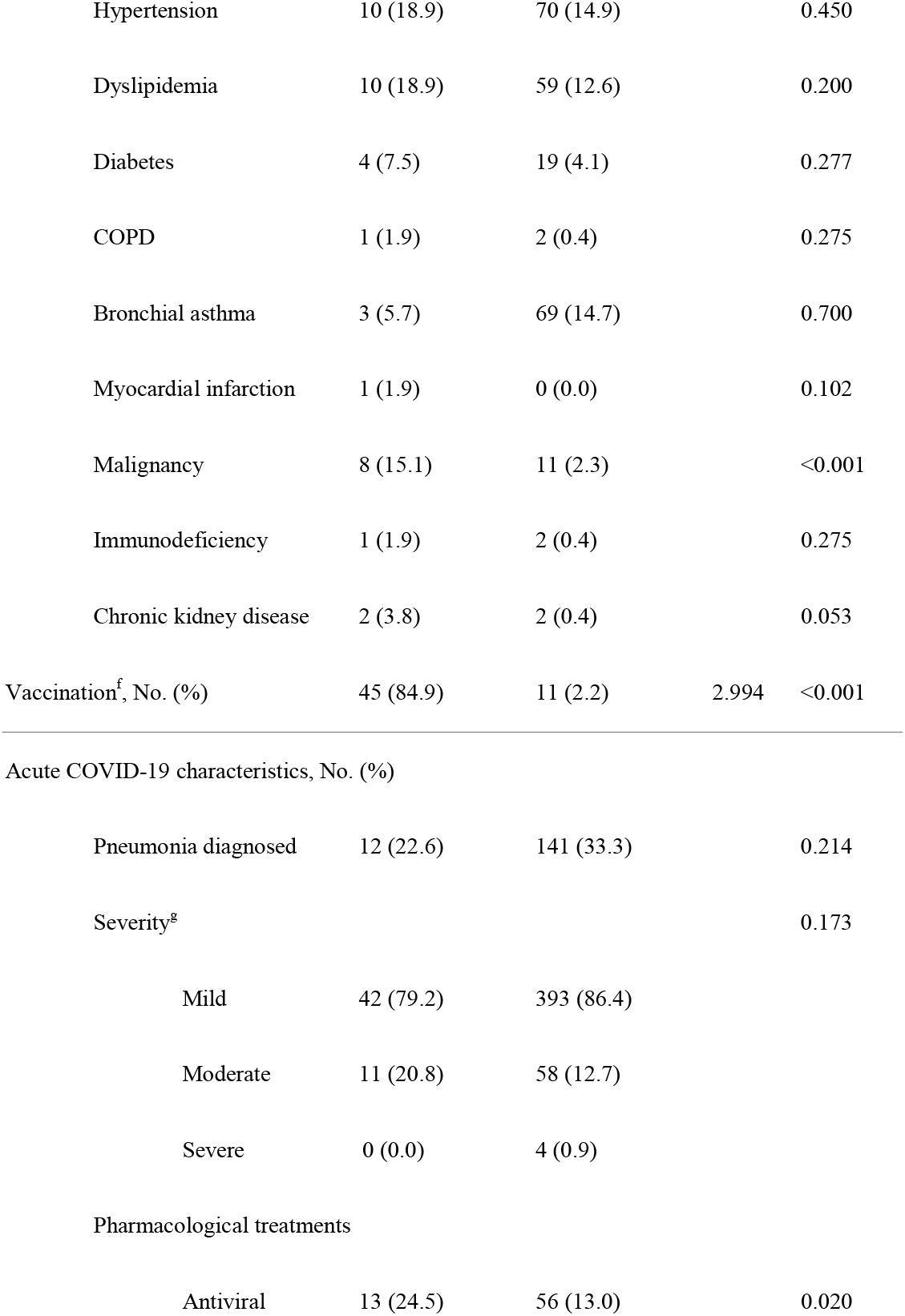
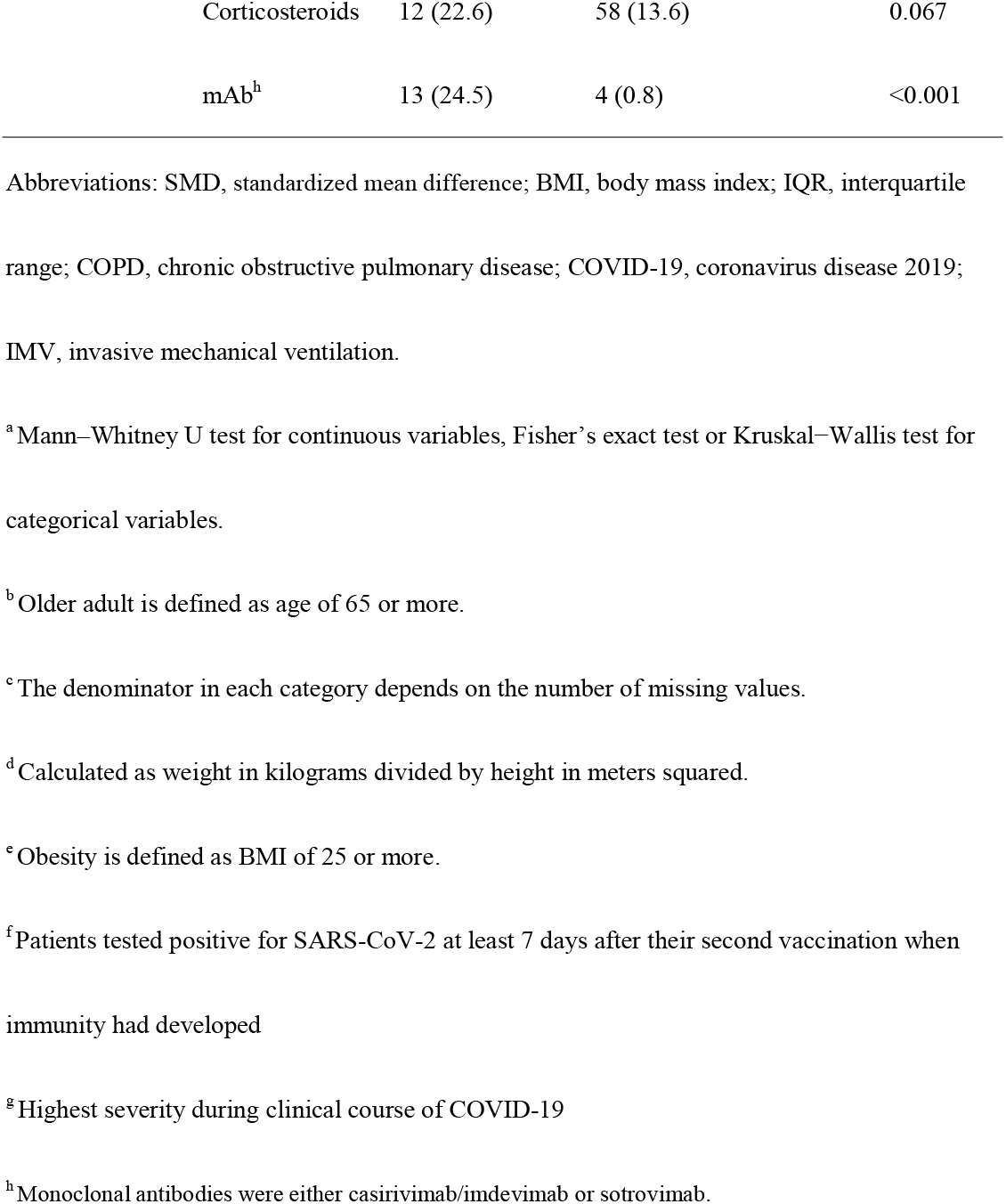
Characteristics of the participants in the Omicron group and the control group before matching

**Table 2.**
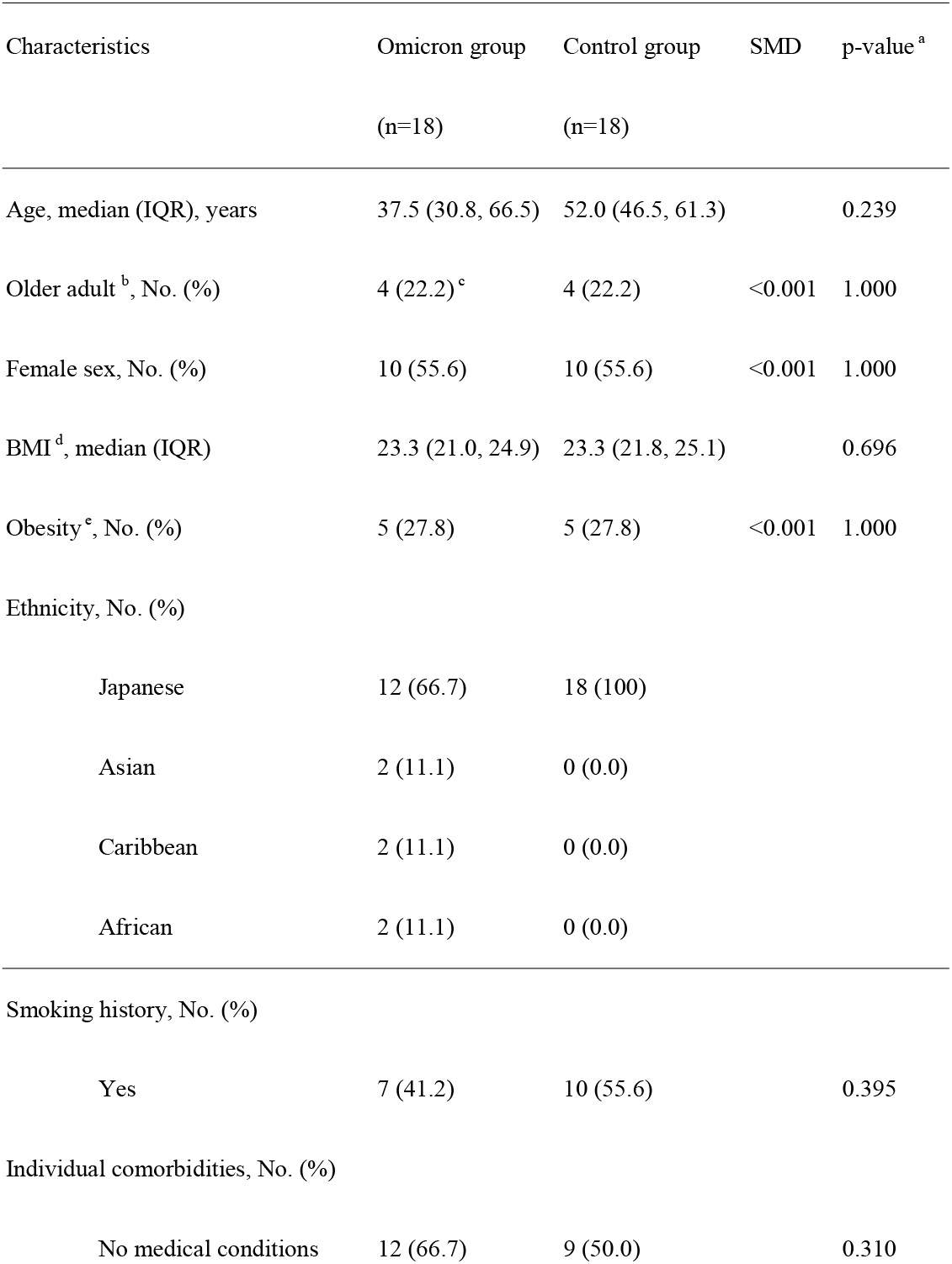
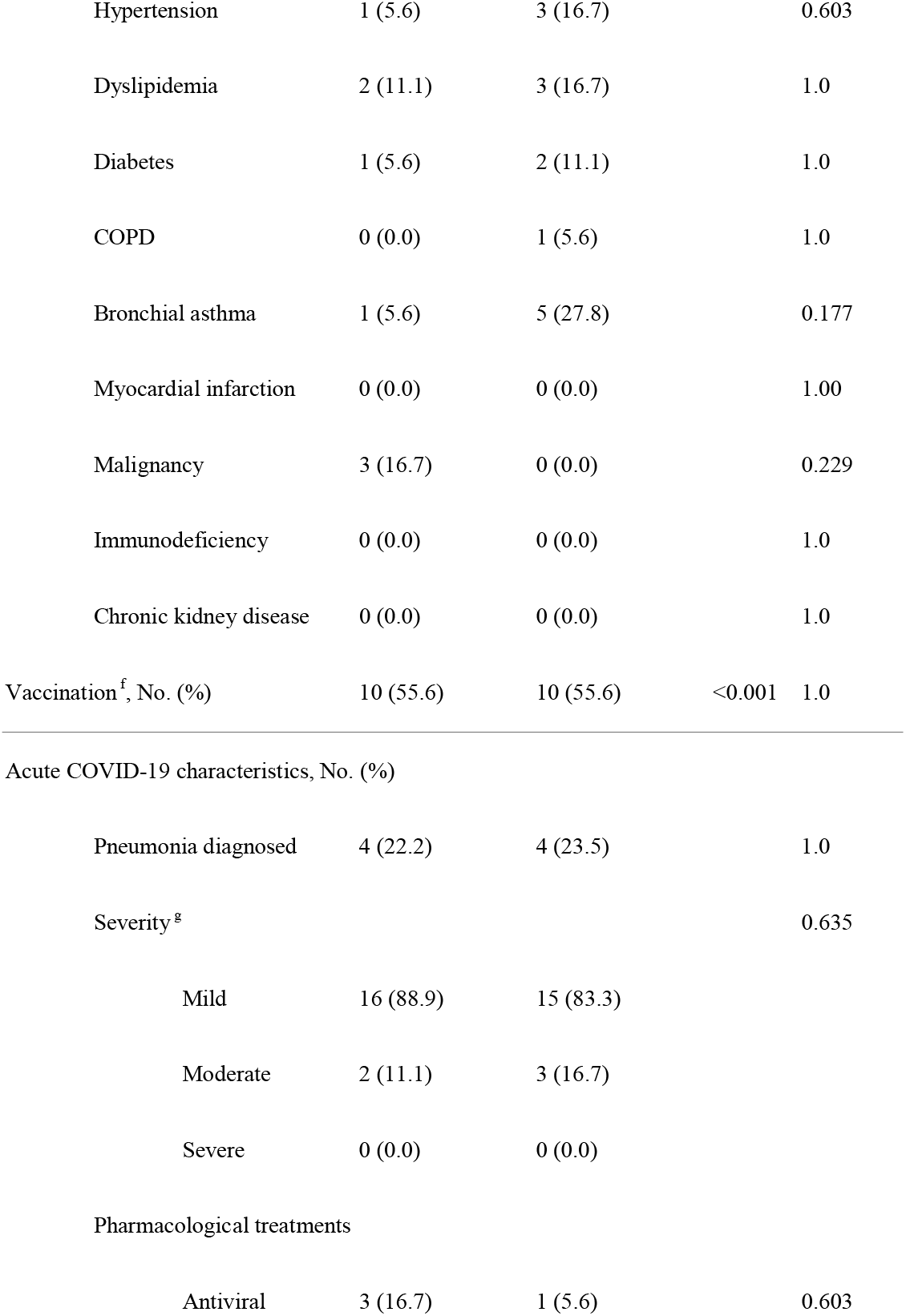
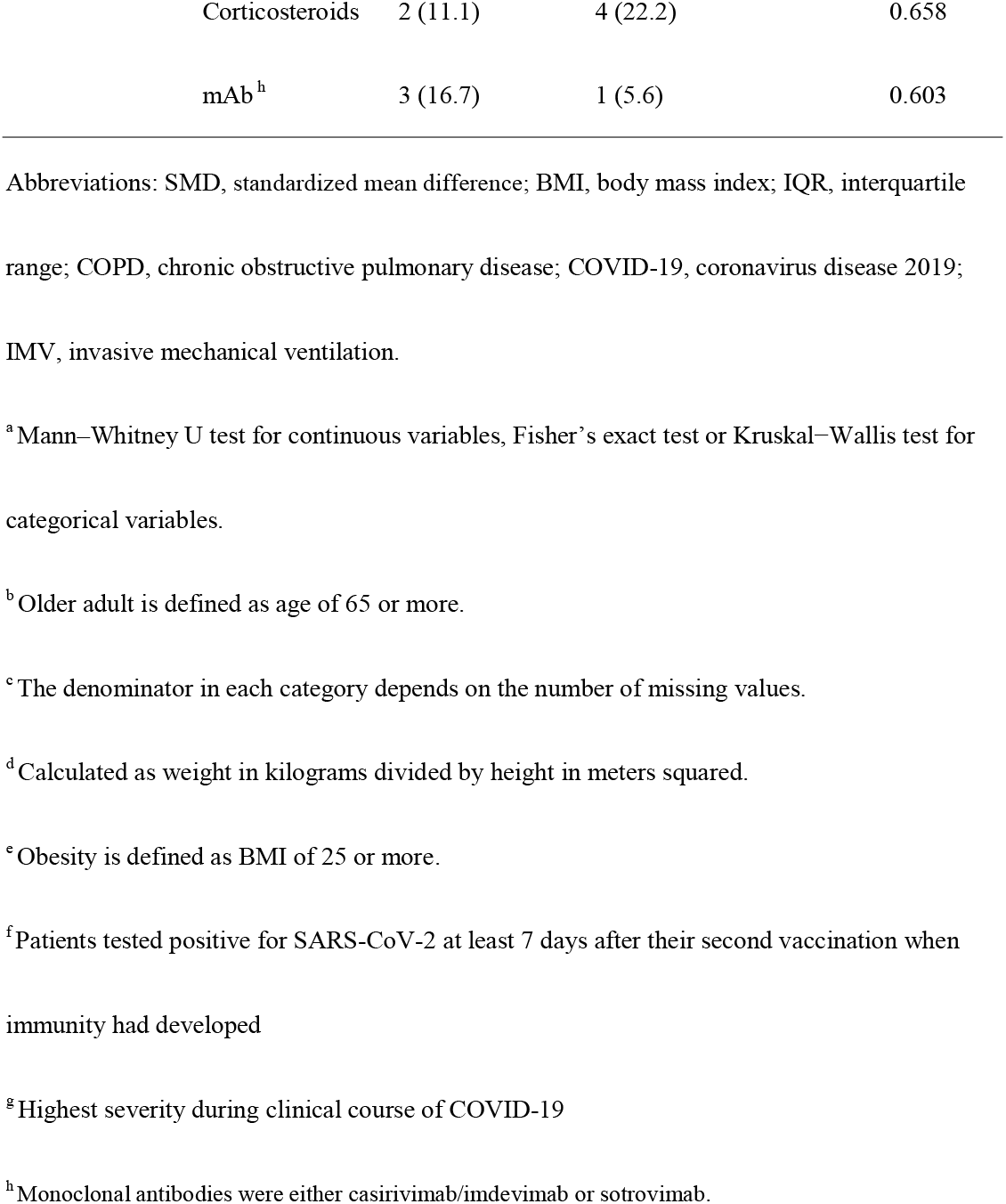
Characteristics of the participants in the Omicron group and the control group after matching

After matching, 18 patients each in the Omicron and control group had improved covariate balance. The number of the older adults (defined as age of 65 and above), female, obese (defined as BMI ≥ 25), and vaccinated patients (defined as patients who tested positive for SARS-CoV-2 at least seven days after their second vaccination) were 4 (22.2%) (standardized mean difference: SMD <0.001, p=1.000), 10 (55.6%) (SMD <0.001, p=1.000), 5 (27.8%) (SMD <0.001, p=1.000), and 10 (55.6%) (SMD <0.001, p=1.000), respectively for both the Omicron and the control group.

### Prevalence of post COVID-19 condition in the Omicron group and the control group

The prevalence of post COVID-19 condition in the Omicron and the control group before matching is summarized in **Table 3** and **Appendix 3**, respectively. In the Omicron group, 3 (5.7%) patients suffered from fatigue, 2 (3.8%) from shortness of breath, 2 (3.8%) from cough, 2 (3.8%) from depressed mood, and 3 patients (5.7%) from loss of concentration that lasted more than 2 months within 3 months since the disease onset. As for the other symptoms of the Omicron group, 45 (84.9%) patients had fever, 30 (56.6%) had sore throat, and 15 (28.3%) had runny nose in the acute phase. However, none of the symptoms lasted more than 2 months within 3 months since the disease onset. In addition, 3 patients (5.7%) had dysosmia, and 7 (13.2%) had dysgeusia.

**Table 3.**
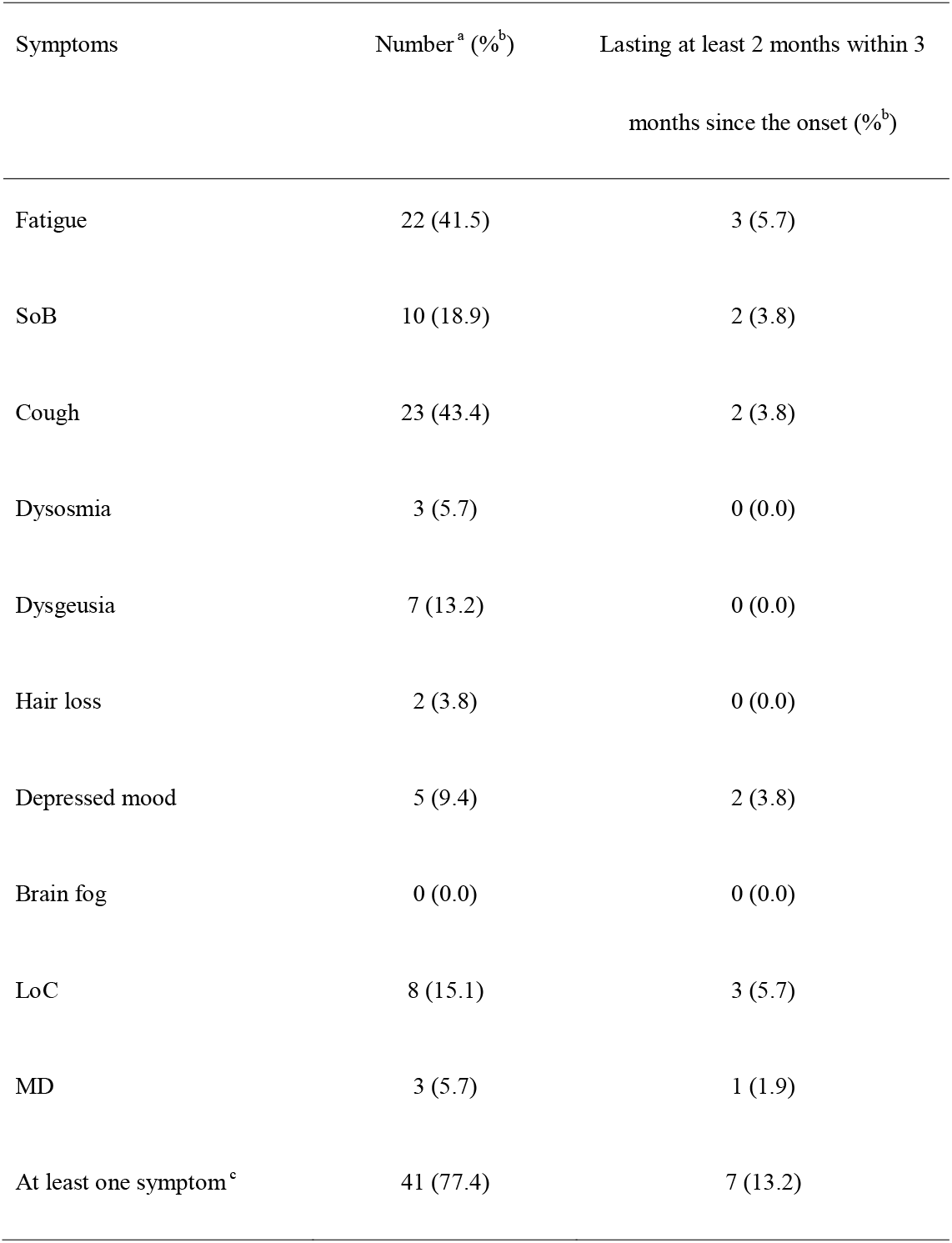
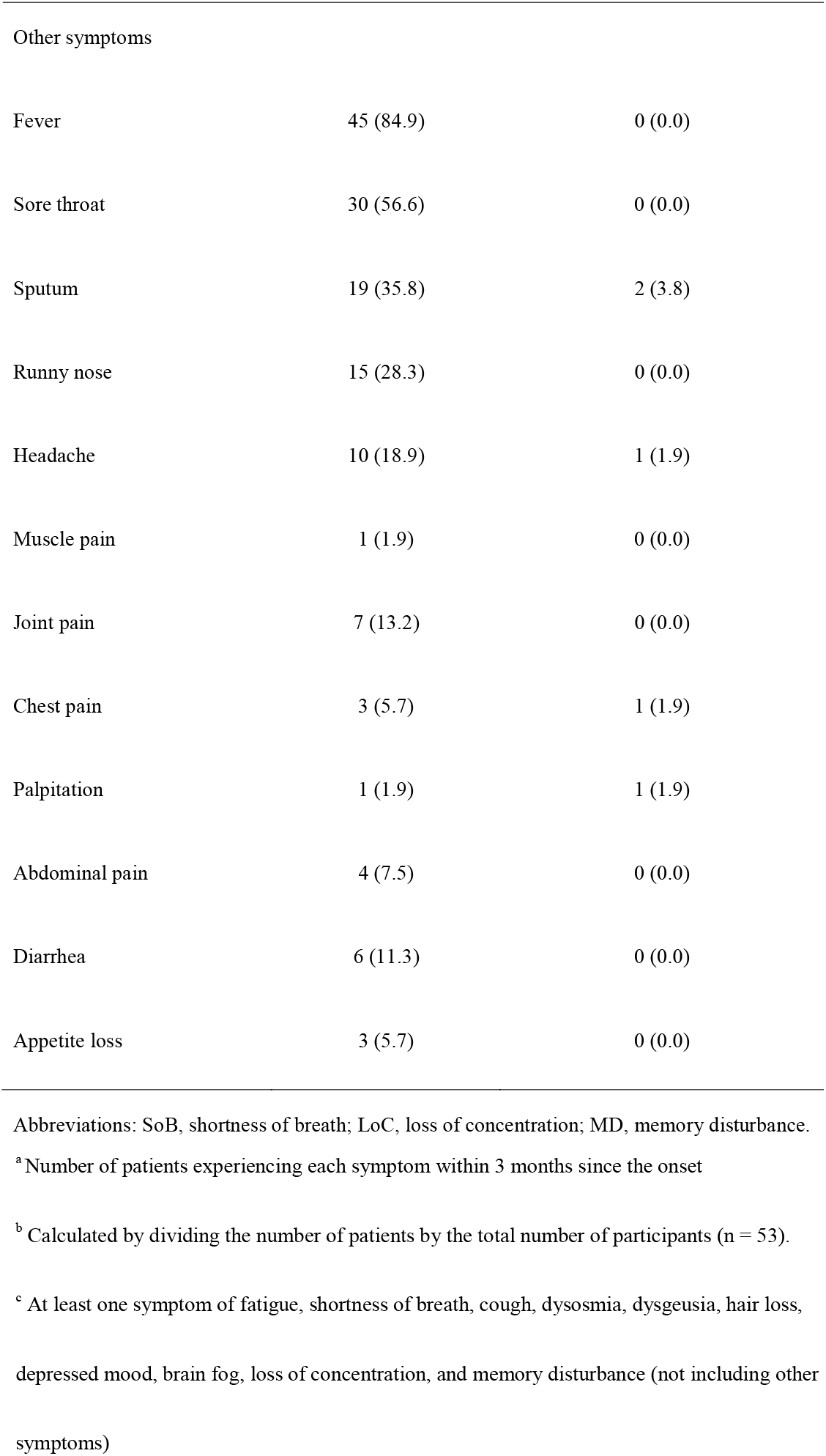
The number of participants with post COVID-19 condition in the Omicron group

| Symptoms | Number <sup>a</sup> (%<sup>b</sup>) | Lasting at least 2 months within 3<br>months since the onset (%<sup>b</sup>) |
| --- | --- | --- |
| Fatigue | 22 (41.5) | 3 (5.7) |
| SoB | 10 (18.9) | 2 (3.8) |
| Cough | 23 (43.4) | 2 (3.8) |
| Dysosmia | 3 (5.7) | 0 (0.0) |
| Dysgeusia | 7 (13.2) | 0 (0.0) |
| Hair loss | 2 (3.8) | 0 (0.0) |
| Depressed mood | 5 (9.4) | 2 (3.8) |
| Brain fog | 0 (0.0) | 0 (0.0) |
| LoC | 8 (15.1) | 3 (5.7) |
| MD | 3 (5.7) | 1 (1.9) |
| At least one symptom <sup>c</sup> | 41 (77.4) | 7 (13.2) |
| Other symptoms |  |  |
| Fever | 45 (84.9) | 0 (0.0) |
| Sore throat | 30 (56.6) | 0 (0.0) |
| Sputum | 19 (35.8) | 2 (3.8) |
| Runny nose | 15 (28.3) | 0 (0.0) |
| Headache | 10 (18.9) | 1 (1.9) |
| Muscle pain | 1 (1.9) | 0 (0.0) |
| Joint pain | 7 (13.2) | 0 (0.0) |
| Chest pain | 3 (5.7) | 1 (1.9) |
| Palpitation | 1 (1.9) | 1 (1.9) |
| Abdominal pain | 4 (7.5) | 0 (0.0) |
| Diarrhea | 6 (11.3) | 0 (0.0) |
| Appetite loss | 3 (5.7) | 0 (0.0) |

Abbreviations: SoB, shortness of breath; LoC, loss of concentration; MD, memory disturbance.
<sup>a</sup> Number of patients experiencing each symptom within 3 months since the onset
<sup>b</sup> Calculated by dividing the number of patients by the total number of participants (n = 53).
<sup>c</sup> At least one symptom of fatigue, shortness of breath, cough, dysosmia, dysgeusia, hair loss,
depressed mood, brain fog, loss of concentration, and memory disturbance (not including other
symptoms)

The prevalence of post COVID-19 condition in the Omicron and the control group after matching is summarized in **Table 4**. There were no significant differences in the prevalence of each post-acute COVID-19 symptom between the two groups. The number of patients with at least one post-acute COVID-19 symptoms in the Omicron and the control group were 1 (5.6%) and 10 (55.6%) (p=0.003), respectively. The frequency and duration of each symptom in the Omicron and the control group are shown in **Appendix 4** and **Appendix 5**, respectively. The frequency and duration of at least one symptom in both groups are shown in **Figure 1**. The number of patients with at least one symptom at 60 and 90 days after symptom onset or diagnosis of COVID-19 in the Omicron group were 1 (5.6%) and 1 (5.6%), respectively. Meanwhile, 12 (66.7%) and 7 (38.9%) patients in the control group had at least one symptom that lasted at 60 and 90 days, respectively.

**Table 4.**
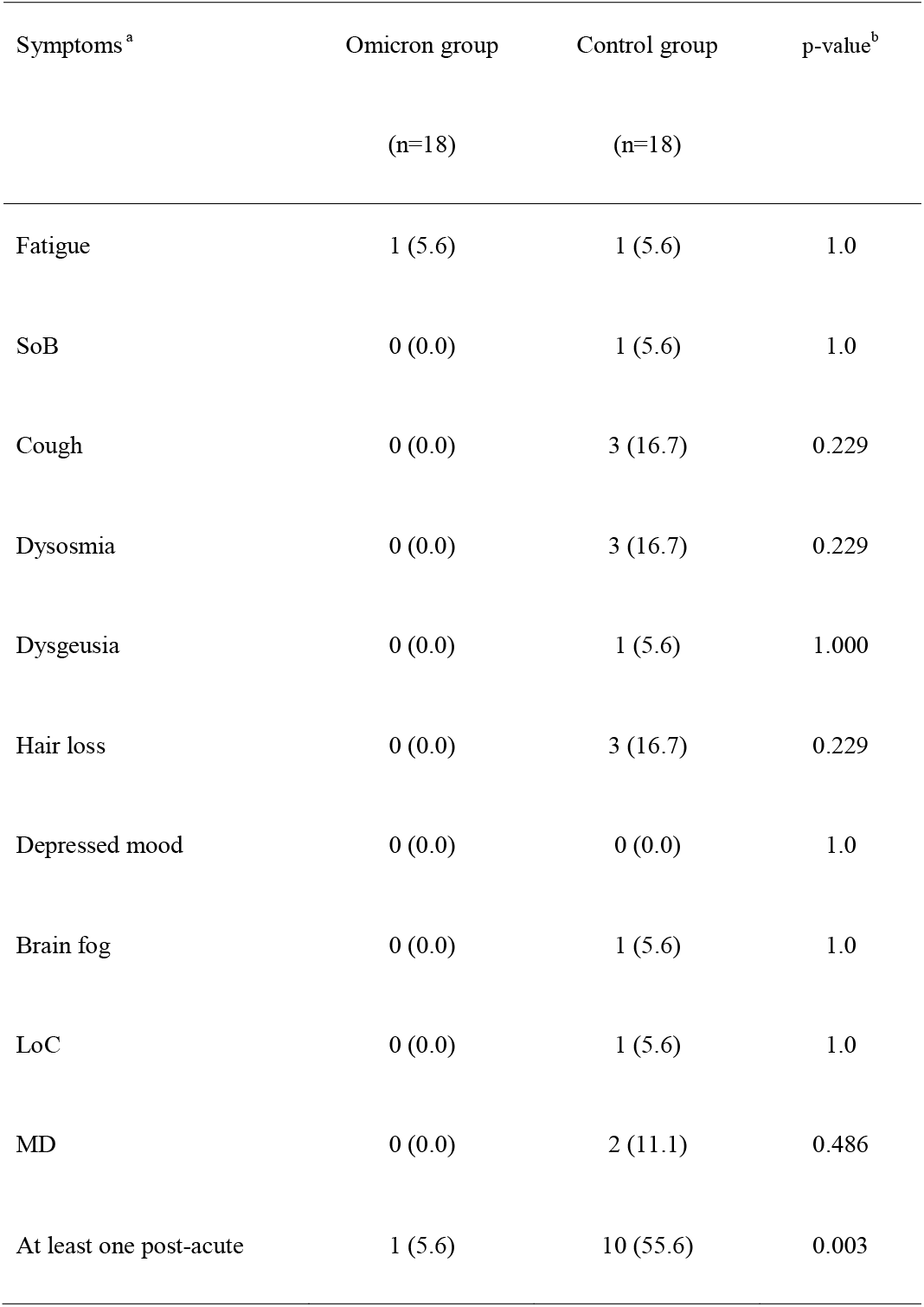
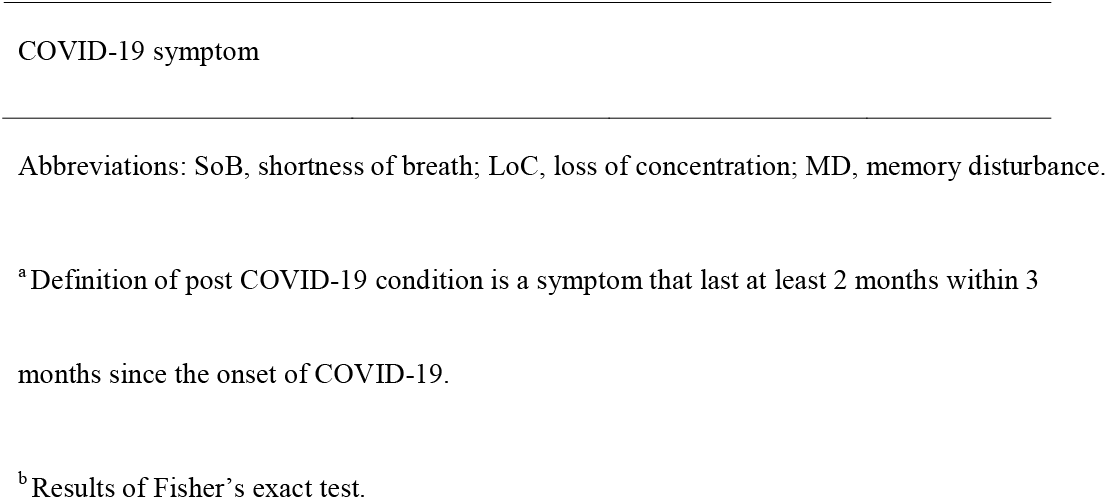
Prevalence of post COVID-19 condition in the Omicron group and the control group

| Symptoms<sup>a</sup> | Omicron group<br>(n=18) | Control group<br>(n=18) | p-value<sup>b</sup> |
| --- | --- | --- | --- |
| Fatigue | 1 (5.6) | 1 (5.6) | 1.0 |
| SoB | 0 (0.0) | 1 (5.6) | 1.0 |
| Cough | 0 (0.0) | 3 (16.7) | 0.229 |
| Dysosmia | 0 (0.0) | 3 (16.7) | 0.229 |
| Dysgeusia | 0 (0.0) | 1 (5.6) | 1.000 |
| Hair loss | 0 (0.0) | 3 (16.7) | 0.229 |
| Depressed mood | 0 (0.0) | 0 (0.0) | 1.0 |
| Brain fog | 0 (0.0) | 1 (5.6) | 1.0 |
| LoC | 0 (0.0) | 1 (5.6) | 1.0 |
| MD | 0 (0.0) | 2 (11.1) | 0.486 |
| At least one post-acute | 1 (5.6) | 10 (55.6) | 0.003 |

Abbreviations: SoB, shortness of breath; LoC, loss of concentration; MD, memory disturbance.
<sup>a</sup> Definition of post COVID-19 condition is a symptom that last at least 2 months within 3
months since the onset of COVID-19.
<sup>b</sup> Results of Fisher's exact test.

**Figure 1.**
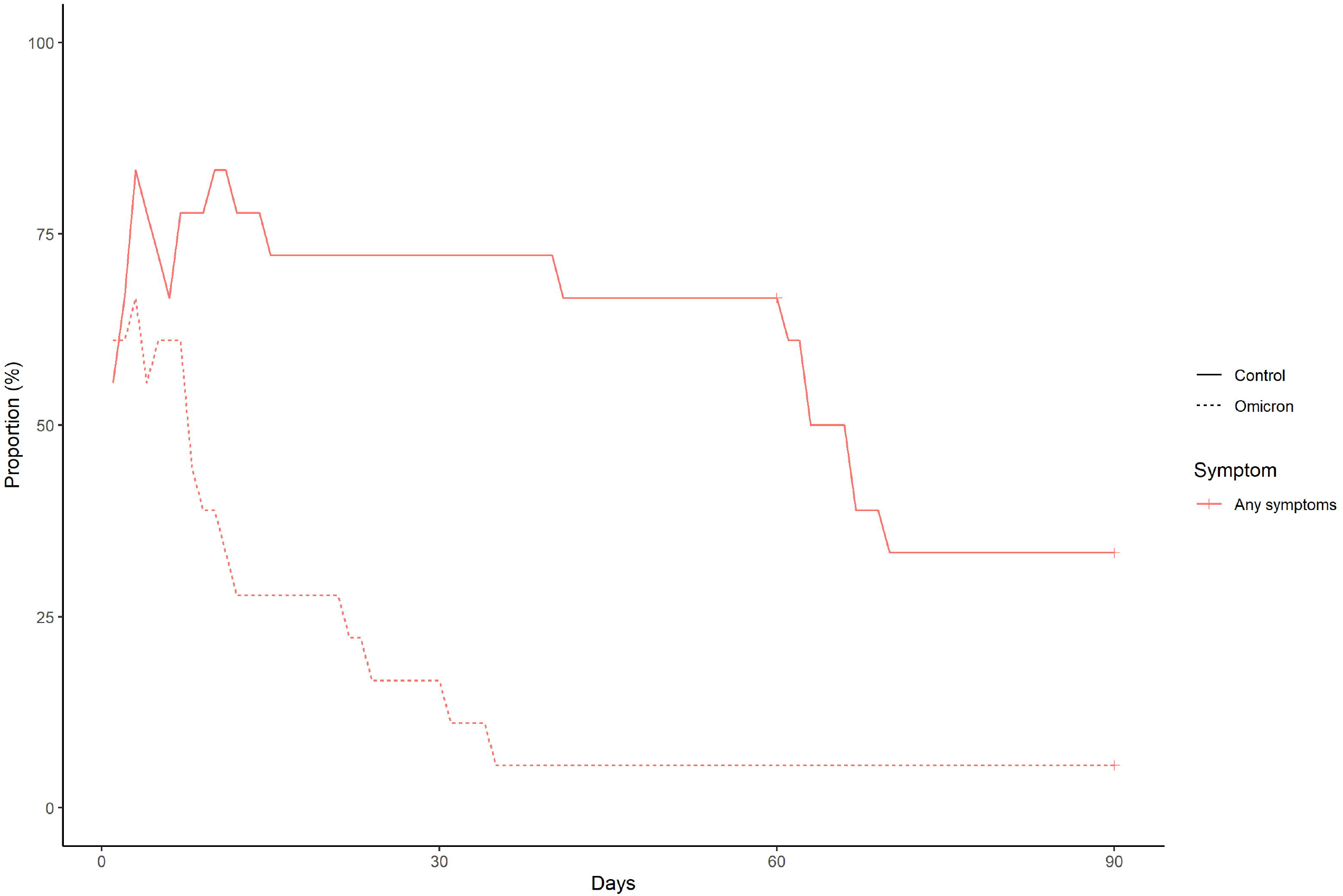
The frequency and duration of at least one symptom in the Omicron group and the control group after matching.

## Discussion

In this study, we investigated and compared the prevalence of post COVID-19 condition in patients with Omicron variant and the previous strains after performing PSM. Since we matched on age, sex, BMI and vaccination status, these factors are unlikely to confound our observation on the prevalence of post COVID-19 condition [17, 18]. The main finding of this study was that there were no significant differences in the prevalence of post COVID-19 conditions between the Omicron and the control group although the prevalence in the Omicron group tended to be less than that in the control group. In addition, the number of patients with at least one post-acute COVID-19 symptom in the Omicron group was significantly less than that in the control group. However, several patients were infected with Omicron variants worldwide. In particular, World Health Organization reported that among the 432,470 specimens collected from 13 January to 11 February 2022, 98.3% were Omicron variant [9]. Thus, while the prevalence rate may be lower in Omicron group, the actual number of patients suffering from post COVID-19 condition can be high. Further research with several participants is needed to investigate the precise epidemiology of post COVID-19 conditions due to Omicron, and assess its impact on health-related quality of life and social productivity.

We found that 84.9%, 56.6%, and 28.3% of patients with Omicron had fever, sore throat, and runny nose, respectively, but none of which lasted more than 2 months within 3 months since the onset of the disease. Cristina Menni et al. reported that sore throat was more common with Omicron than with Delta variant (70.5% *vs* 60.8%, odds ratio 1.55; 95% CI 1.43–1.69, p<0.001) [7], indicating that patients with Omicron tended to have upper respiratory symptoms. This is also consistent with an *in vitro* study stating that Omicron replicates faster than all other SARS-CoV-2 variants in the bronchus but less efficiently in the lung parenchyma, and appears to enter human cells by a different route [22]. Although we could not compare the prevalence and duration of these symptoms of Omicron with those of previous strains, characterizing the clinical course of infection by Omicron is not only of direct public health relevance providing public and clinicians awareness on what symptoms to look out for, but will also assist in understanding the potential effects of future variants of concern [7].

We also found that 5.7%, 13.2% and 3.8% of the patients with Omicron had dysosmia, dysgeusia, and hair loss, respectively. This finding was consistent with a previous report stating that loss of smell was less common in participants infected with Omicron than with Delta variant (16.7% *vs* 52.7%, odds ratio 0.17; 95% CI 0.16–0.19, p<0.001) [7]. These findings can also help raise awareness on what symptoms are more common with the Omicron variant to that can aide in rapid diagnosis, minimizing the spread of COVID-19.

Our study had several limitations. First, we compared data obtained from two surveys with different methodologies: data of Omicron group from telephone interviews, and those of control group from a self-reported questionnaire-based online/paper-based survey, which may have influenced the results. Second, those with more severe symptoms might less likely to respond to telephone calls if they were subsequently hospitalized, making data on the critically ill more likely to be scarce. Third, the self-reported questionnaire-based online/paper-based survey was subject to various biases, such as selection, volunteer, and recall biases. In particular, the survey was limited to COVID-19 convalescent plasmapheresis patients who underwent the pre-donation screening test. It is unclear whether the results of this survey can be applied to all patients recovering from COVID-19. Forth, this was a single-center study with a small sample size. Fifth, the definition of post COVID-19 condition in this study was similar to a previous study [3]. However, whether the symptoms could be explained by an alternative diagnosis remains unclear in our study. Therefore, our findings may have overestimated the prevalence of post COVID-19 condition. Sixth, some patients had persistent symptoms at the time of the survey. In these cases, the actual duration of the symptoms was unclear. Long-term observation is needed to better understand the duration of post-COVID conditions of Omicron. Seventh, it was impossible to determine the type of variant in the control group. Considering that 31.5%, 48.6%, and 19.9% in the control group got infected with COVID-19 between February 2020 and October 2020, November 2020 and June 2021, and July 2021 and October 2021, respectively, it was likely that alpha (B.1.1.7) strains were most predominant [25][26][27]. The prevalence of the SARS-CoV-2 variants may have influenced the frequency of post-COVID conditions in the control group. Lastly, we could not identify each variant in Omicron (B.1.1.529, BA.1, BA.1.1, BA.2, BA.3, BA.4 and BA.5 lineages).

## Conclusion

We investigated the prevalence of post COVID-19 conditions in patients with Omicron and other strains of SARS-CoV-2 after matching on age, sex, BMI, and vaccination status. There were no significant differences in the prevalence of each post-acute COVID-19 symptom between the Omicron and the control group. However, the number of patients with at least one post-acute COVID-19 symptom in the Omicron group was significantly less than that in the control group. As many patients worldwide were infected with Omicron, several patients may suffer from post-acute COVID-19 symptoms. Further research with more participants is needed to investigate more precise epidemiology of post COVID-19 condition due to Omicron, and its impact on health-related quality of life and social productivity.

## Data Availability

All data produced in the present work are contained in the manuscript.

## Acknowledgments

We thank all the people who participated in our study.

## Figure Legends

**Appendix 4**. The frequency and duration of each symptom in the Omicron group

**Appendix 5**. The frequency and duration of each symptom in the control group

