## Appendix 1 for "Post COVID-19 condition of the Omicron variant of SARS-CoV-2"

Interview Guide

1. Introduce yourself and inform that you are a member of the COVID-19 medical team in National Center for Global Health and Medicine. After verifying the patient identity, explain the purpose and the outline of the telephone interview survey, and obtain consent in accordance with the contents of the research protocol. Conduct below interview if patient agreed.
2. Check the patient’s medical record. Then, explain the following and proceed with the interview.

- When you were admitted to our hospital, it was recorded that you had symptoms of ____. Are you sure that you had those symptoms?

- Some of the data regarding your symptoms are missing. Do you have any of the following (fever, malaise, dyspnea, arthralgia, myalgia, chest pain, cough, abdominal pain, olfactory disturbance, gustatory disturbance, nasal discharge, headache, sputum, sore throat, diarrhea, nausea, vomiting, anorexia, hair loss, depression, impaired concentration, impaired memory)?

- Is it correct to say that you had symptoms of ________________ and ________________ during your medical treatment for COVID-19?

- How many weeks or months did the symptoms persist? When did you notice improvement? (If you don't know, were you symptomatic at one, two or three months after onset?)

- Were there any new symptoms that appeared after your recovery? (If unclear, were there any symptoms at 1 or 2 months after onset?

3. If interviewee has medical questions other than those in this survey, listen to the patient and connect with Dr. Morioka, the principal investigator.
