## Appendix 2 for "Post COVID-19 condition of the Omicron variant of SARS-CoV-2"

If you agree to participate in this survey, please check the box below.

- I agree to participate in this study.

　We would like to ask you some questions about yourself. Please check all that apply and fill in the information in parentheses.

Age　	（　　　　　）

Sex　	1. Man　　2. Woman　　3. other

Height　（　　　　　）cm

Weight	（　　　　　）kg

Smoking history 1. currently smoking 2. smoked in the past 3. have never smoked at all

Drinking history	1. yes 2. No

Medical history

1. hypertension 2. diabetes 3. dyslipidemia 4. bronchial asthma 5. COPD (Chronic obstructive pulmonary disease) 6. myocardial infarction 7. malignant tumor 8. rheumatoid collagen disease 9. immunodeficiency disease 10. chronic kidney disease 11. Neurological diseases such as Parkinson's disease or Alzheimer's disease 12. other (	)

If any of the above 7, 8, 9, or 11 apply to you, what is the specific name for your illness?（　　　　　　　　　　　　　）

Pregnancy history (female only, multiple answers possible)

1. Had a pregnancy before the new coronavirus infection

2. Was pregnant at the time of the new coronavirus infection

3. Became pregnant after the new coronavirus infection

4. Have never been pregnant

②　I would like to ask you about your coronavirus infection.

　(If you answered this question in the previous survey in April 2021, you do not need to fill it in.)

When did you first become aware of your symptoms?

(	)

When were you first diagnosed with the new coronavirus?

Please respond to all of the following

Diagnosed as pneumonia?	1. yes 2. no 3. don't know

If yes, when were you diagnosed with pneumonia? (	) ( year, month, day)

Administered oxygen at least once	1. yes 2. no 3. don't know

Treated with a ventilator at least once	1. yes 2. no 3. don't know

Treated with extracorporeal membrane oxygenation (ECMO) at least once 1. yes 2. no 3. don't know

Received antiviral medication	1. yes 2. no 3. don't know

Received steroids:	1. yes 2. no 3. don't know

Received antibody cocktail? (casirivimab and imdevimab)

1. yes	2. No	3. Don’t know

Received antibody? (Sotrovimab)

1. yes	2. No	3. Don’t know

If yes, when did you receive the antibody?　　(year/month/day	)

Vaccination history for new coronavirus

1. vaccinated only once 2. vaccinated twice 3. vaccinated three times 4. not vaccinated

If vaccinated, please select the type of vaccine you received from the following list.

1. Pfizer (BioNTech) 2. Moderna 3. AstraZeneca 4. Other (	)

Type of vaccine (	) Date of vaccination ( year/month/ day )

Type of vaccine (	) Date of inoculation ( year/month/day)

Type of vaccine (	) Date of inoculation ( year month day )

　We would like to ask you about the symptoms of the new coronavirus infection. Please indicate whether you have any of the following symptoms, and if so, for how many days?

(Example) If the symptom started 30 days after the disease onset, and lasted for 20 days, please answer as follows.

①. yes	2. no

If yes, please answer as follows:

A. From the time of onset	B. From day ( 30 ) after onset

a. Lasted for (20) days	b. Continues to this day

Fatigue (sluggishness of the body)

1. yes	2. no

If yes, please answer as follows:

A. From the time of onset	B. From day (	) after onset

a. Lasted for (	) days	b. Continues to this day

Dyspnea (difficulty breathing)

1. yes	2. no

If yes, please answer as follows:

A. From the time of onset	B. From day (	) after onset

a. Lasted for (	) days	b. Continues to this day

Cough

1. yes	2. no

If yes, please answer as follows:

A. From the time of onset	B. From day (	) after onset

a. Lasted for (	) days	b. Continues to this day

Olfactory impairment (difficulty in perceiving smells)

1. yes	2. no

If yes, please answer as follows:

A. From the time of onset	B. from day (	) after onset

a. Lasted for (	) days	b. Continues to this day

Taste disorder (difficulty in tasting food)

1. yes	2. no

If yes, please answer as follows:

A. From the time of onset	B. From day (	) after onset

a. Lasted for (	) days	b. Continues to this day

Hair loss

1. yes	2. no

If yes, please answer as follows:

A. From the time of onset	B. From day (	) after onset

a. Lasted for (	) days	b. Continues to this day

Depression (feeling down)

1. yes	2. no

If yes, please answer as follows:

A. From the time of onset	B. From day (	) after onset

a. Lasted for (	) days	b. Continues to this day

Lightheadedness

1. yes	2. no

If yes, please answer as follows:

A. From the time of onset	B. From day (	) after onset

a. Lasted for (	) days	b. Continues to this day

Difficulty concentrating

1. yes	2. no

If yes, please answer as follows:

A. From the time of onset	B. From day (	) after onset

a. Lasted for (	) days	b. Continues to this day

Forgetfulness

1. yes	2. no

If yes, please answer as follows:

A. From the time of onset	B. From day (	) after onset

a. Lasted for (	) days	b. Continues to this day
