## Appendix 3 for "Post COVID-19 condition of the Omicron variant of SARS-CoV-2"

**Appendix 3.** The number of participants with post COVID-19 condition in the control group (n=502)

| Symptoms | Number ^a^ (%^b^) | Lasting at least 2 months within 3 months since the onset (%^b^) |
| --- | --- | --- |
| Fatigue | 394 (78.5) | 61 (12.2) |
| SoB | 183 (36.5) | 39 (7.8) |
| Cough | 299 (59.6) | 28 (5.6) |
| Dysosmia | 290 (57.8) | 84 (16.7) |
| Dysgeusia | 242 (48.2) | 51 (10.2) |
| Hair loss | 147 (29.3) | 51 (10.2) |
| Depressed mood | 147 (29.3) | 58 (11.6) |
| Brain fog | 186 (37.1) | 74 (14.7) |
| LoC | 185 (36.9) | 84 (16.7) |
| MD | 110 (21.9) | 69 (13.7) |
| At least one symptom | 481 (95.8) | 212 (42.2) |

Abbreviations: SoB, shortness of breath; LoC, loss of concentration; MD, memory disturbance.

^a^ Number of patients experiencing each symptom within 3 months since the onset

^b^ Calculated by dividing the number of patients by the total number of participants (n = 502).
