## Supplementary figures and images for "Post COVID-19 condition of the Omicron variant of SARS-CoV-2"

### Appendix 4

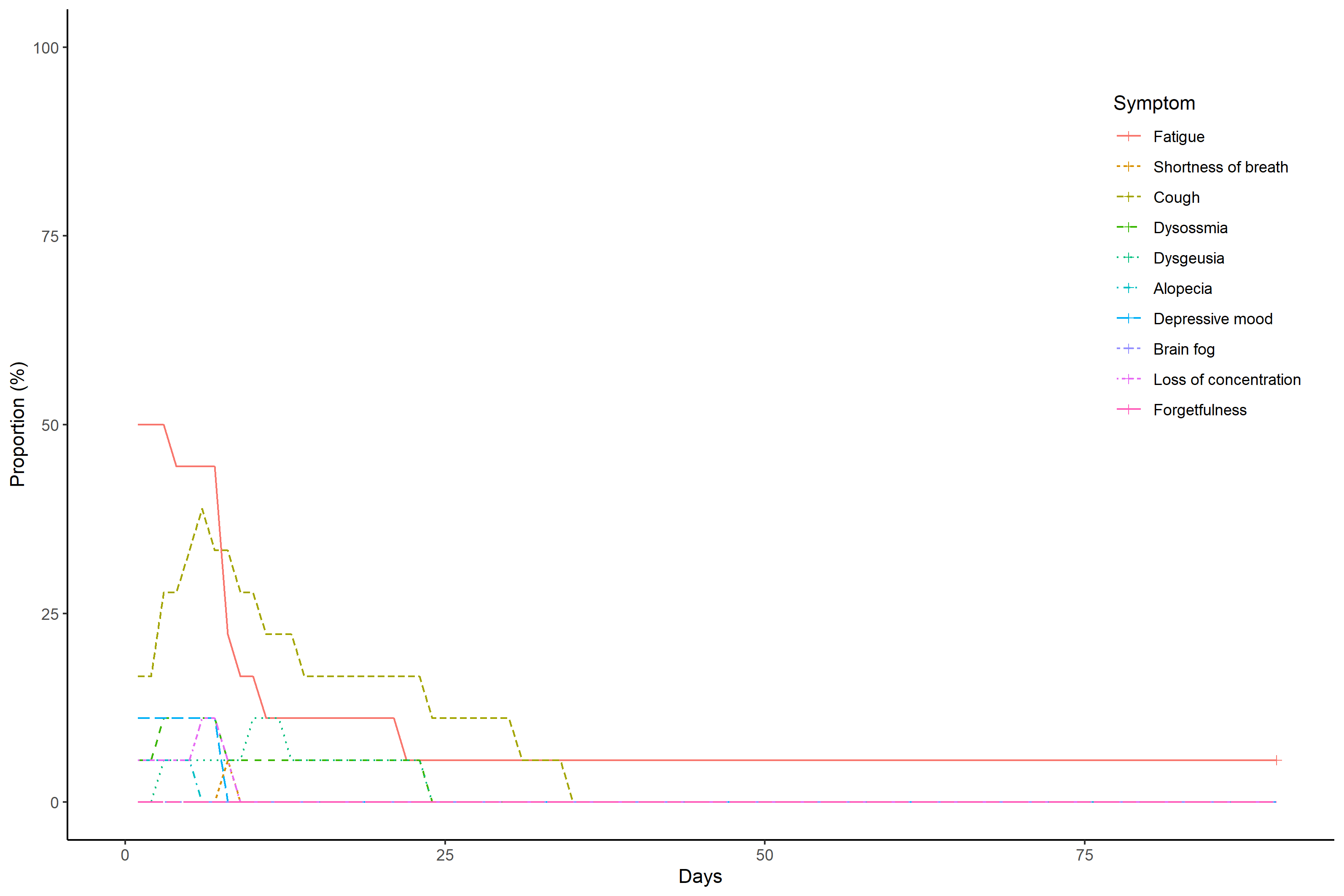

### Appendix 5

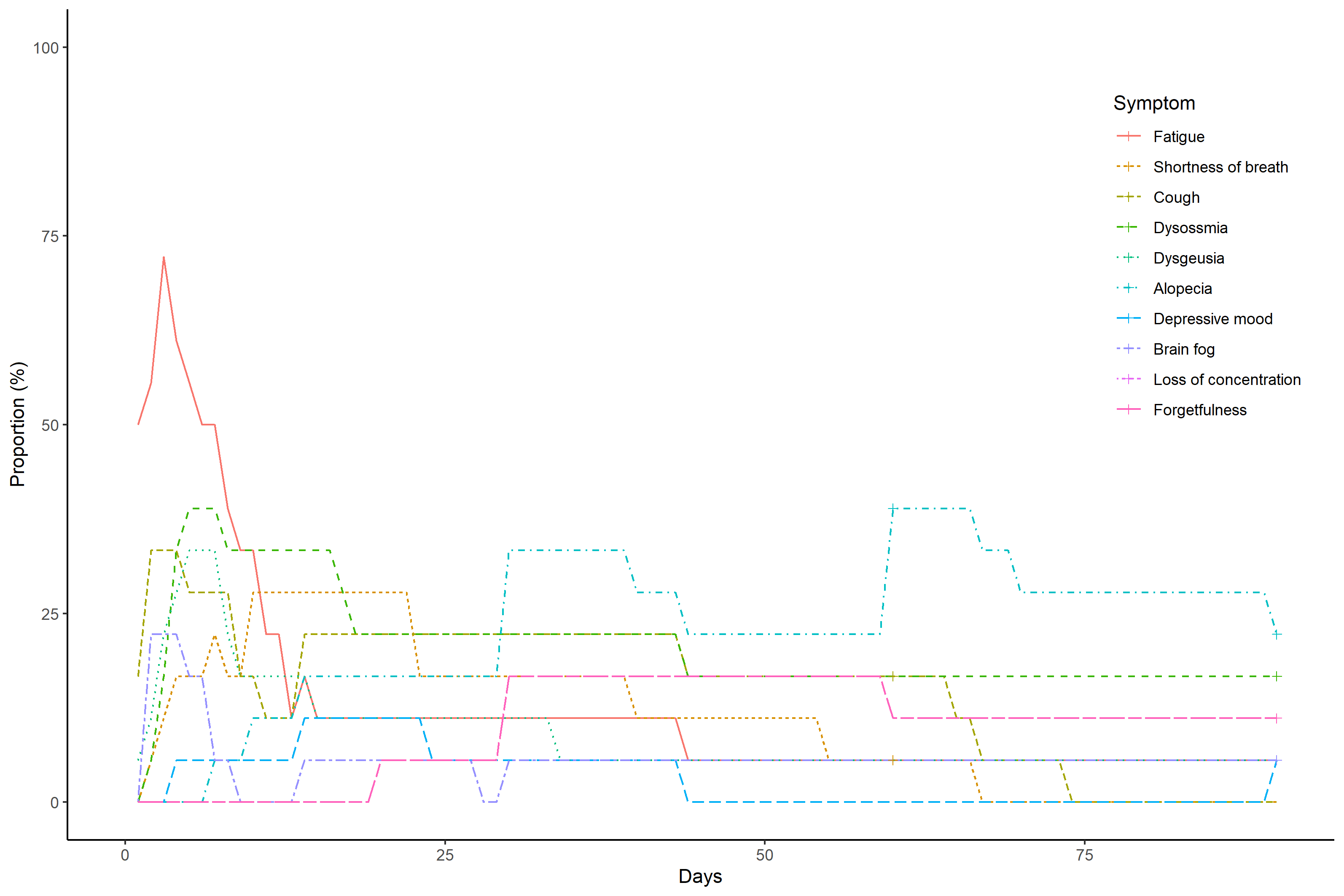
